# P. acnes qPCR-based antibiotics resistance assay (ACQUIRE) reveals widespread macrolide resistance in acne patients and can eliminate macrolide misuse in acne treatment

**DOI:** 10.1101/2021.06.21.21256023

**Authors:** Jingheng Zhang, Fang Yu, Keyun Fu, Xinyu Ma, Yi Han, Chi Ching Ali, Haonan Zhou, Yantao Xu, Tingyue Zhang, Shuntong Kang, Yiming Xu, Zhuolin Li, Jiaqi Shi, Shuai Gao, Yongyi Chen, Jianglin Zhang, Liyu Chen, Feizhou Zhu

## Abstract

**Background:** Macrolides have been widely used to treat moderate-to-severe acne for more than 50 years. However, prevalent antibiotic resistance of *Propionibacterium acnes*, along with the absence of clinically available resistance tests, has made macrolide misuse a frequent occurrence, with serious consequences.

**Objective:** We developed *P. acnes* qPCR-based antibiotics resistance assay (ACQUIRE) to enable fast and accurate detection of *P. acnes* macrolide resistance in clinical settings, representing an opportunity to administer antibiotics more wisely and improve the quality of care.

**Methods:** A cross-sectional observational study was conducted to probe into the macrolide resistance of *P. acnes* in acne patients.

**Results:** The high sensitivity of ACQUIRE enabled us to reveal a much higher *P. acnes* 23S rDNA point mutation rate (52%) and thus a higher macrolide resistance (75.5%) compared to previous reports. Carriage of ermX gene was discovered on 472 (53%) subjects, which concurs with previous studies.

**Conclusion:** Macrolide resistance of *P. acnes* is much higher than previously reported.

## Body of manuscript

Acne vulgaris, one of the most common skin disorders worldwide, is inflicting billions of patients, significantly impairs their quality of life. The pathogenesis of acne involves the release of inflammatory mediators, skin hyper-keratinization, increased sebum secretion, and colonization of *Propionibacterium acnes* (recently reclassified as *Cutibacterium acne*s)[1-5]. As the dominant commensal of the human pilosebaceous unit, *P. acnes* is intimately involved in the development of acne. It not only disturbs the proliferation of keratinocytes, but also carries lipase, protease, and hyaluronidase activity which can damage the pilosebaceous unit and induce inflammation[6]. *P. acnes* can also activate toll-like receptors(TLRs)[7] and protease-activated receptors expressed by keratinocytes, which in turn induces the production of interleukins(ILs) and matrix metalloproteinases(MMPs)[8]. The production of anti-*p. acnes* antibodies can also activate complement system. Therefore, inhibiting the overgrowth of *P. acnes* has been one of the major themes in acne treatment. Being able to inhibit *P. acnes* and reduce inflammation concurrently, antibiotics have been used in acne treatment for more than 50 years, representing an essential part of the first-line treatment for moderate-to-severe acne[9]. Macrolides (erythromycin, clarithromycin and azithromycin) and tetracyclines (minocycline and doxycycline) are two classes of antibiotics that are frequently applied in acne treatment[1 10 11]. However, compared with macrolide, tetracyclines have several shortcomings, including more common adverse effects, potential liver and kidney toxicity and most importantly, its incompatibility with oral isotretinoin, which made it impossible to start oral antibiotic and oral isotretinoin therapy simultaneously. Hence macrolide remains essential in acne management in China[12], Japan[13] and many other countries[14].

However, the extensive use of macrolides has caused escalating *P. acnes* resistance worldwide. Increases in *P. acnes* resistance have now been reported in all major regions within China and around the globe[12 13 15-22], with many countries reporting more than 50% of macrolide-resistant *P. acnes*[14]. If macrolide is prescribed, the carriage of macrolide-resistant *P. acnes* strains may cause reduced treatment response, relapse, or prolonged course of disease for acne patients, increasing the disease burden of acne.

To worsen the situation, current antimicrobial susceptibility test (AST) of *P. acnes* employs a series of time-, labor- and cost-intensive procedures due to the nature of *P*. acnes. As an anaerobe, *P*. acnes require special culture environment and propagate slowly, so the test result could only be available after a week post sampling. The sensitivity of current culture-based method is also unsatisfactory because the resistant *P*. acnes would be hard or even impossible to isolate if existing only in low-percentage, which is often the case when mutation occurred naturally but hadn’t undergone antibiotic selection. Those drawbacks not only prevent any chance of applying *P*. acnes resistance test clinically, but also impose huge restrictions on related scientific research.

Toward those ends, we developed *P*. acnes qPCR-based antibiotics resistance assay (ACQUIRE), a method that utilizes the comedone (white head or black head) extracted from patients’ follicle based on ARMS-qPCR (Amplification refractory mutation system-quantitative PCR), and is capable of determining the presence of macrolide-resistant *P. acnes* within 3 hours, showing great potential to serve as a routine test for clinical moderate-to-severe acne patients. ARMS-qPCR is widely used as a convenient and cost-saving tool to detect the point mutation and SNPs in nucleic acids[23]. However, it has not been utilized to determine the resistance determinants in *P. acnes* before this study.

With ACQUIRE, we conducted a large-scale cross-sectional investigation to examine the macrolide resistance level of *P. acnes*.

## Methods

### Participants and study design

A total of 915 patients with acne vulgaris were enrolled from December 2017 to January 2020 at both the Department of Dermatology, Xiangya Hospital, Central South University and a skin care institution (Miao’miao-qingfu) to ensure a wide spectrum of patients. Inclusion criteria were: 12-50 years of age. Exclusion criteria were: history of systemic acne treatment within 6 months. All patients provided informed consent and the study was approved by the ethics committee of Xiangya Hospital. Enrolled patients were all sampled and randomly assigned into two sets (Figure 1).

**Figure 1.**
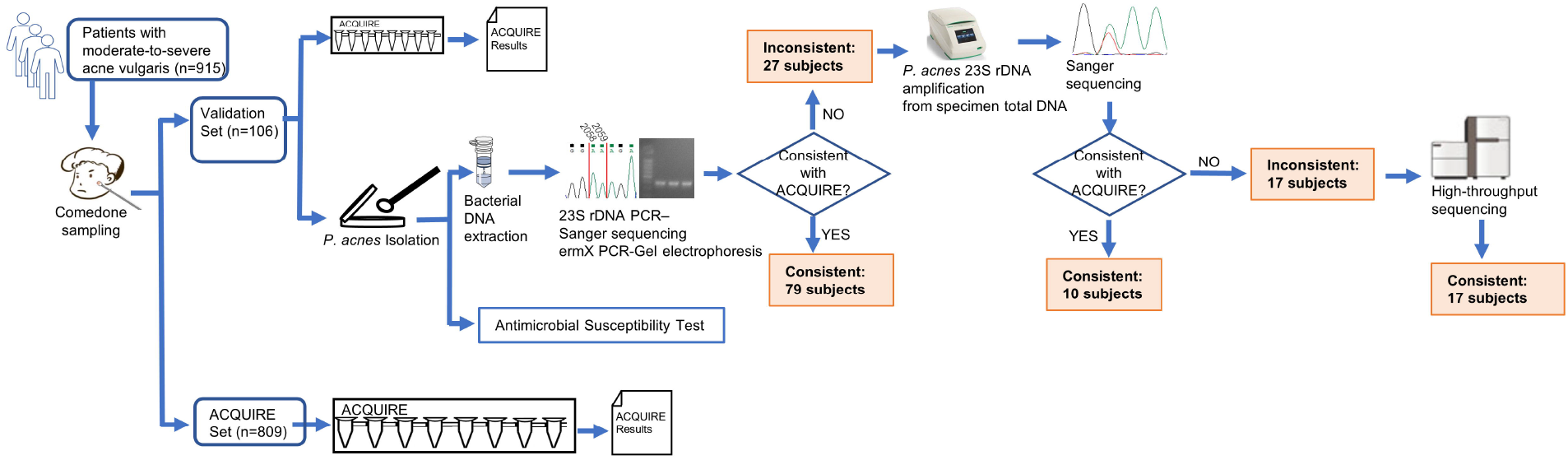
Schematic diagram of study design. Patients enrolled were sampled and randomly assigned into either the ACQUIRE set and the validation set. For patients in Validation Set whose ACQUIRE results and result of culture method were inconsistent, the *P*. acnes 23S rDNA were amplified from specimen total DNA and subject to sanger sequencing. If ACQUIRE results and result of sanger sequencing were inconsistent, the amplification product was subject to high-throughput sequencing (Illumina Miseq), sequence filtration (sequences with less-than-95% similarity to *P*. acnes were discarded) and mutation analysis.

To the validation set (n=106), acne lesions from multiple sites of the subjects’ faces were squeezed using a sterile comedone extractor to obtain the comedone specimens (white head or black head). The specimens were then tested by ACQUIRE. A small portion of the specimens were reserved for *P. acnes* isolation in order to compare the results of ACQUIRE and the current method.

To the ACQUIRE set (n=809), the specimens were also obtained and tested with ACQUIRE.

Specimens were obtained from 915 subjects and tested by ACQUIRE. Subjects were 12–49 years of age (mean ± SD, 24.1±5.7 years). Age and sex distributions among twelve genotypes were not significantly different.

### *P. acnes* isolation, identification, genotyping and antimicrobial susceptibility testing

Isolation, identification, 23S rDNA mutation and ermX detection, and antimicrobial susceptibility test of *P. acnes* were adopted from previous studies[24]. Briefly, *P. acnes* were isolated on CDC anaerobic blood agar under anaerobic atmosphere and 37 °C. The16S rRNA gene of isolated bacteria was amplified and sequenced to identify *P. acnes* from isolated strains. The ermX carriage status and the genotype of 23S rDNA of isolated *P. acnes* were determined by PCR -agarose gel electrophoresis and PCR-sanger sequencing, respectively. The minimum inhibitory concentration (MIC) of *P. acnes* to erythromycin was measured by broth microdilution. A MIC above 2 μ g/mL was considered resistance.

### The development and application of ACQUIRE

Mechanistically, *P. acnes* gains macrolide resistance through the presence of ermX gene in its genome and 23S rDNA point mutation. The ermX gene, which often is carried by a corynebacterial-origin transposon Tn5432, makes *P. acnes* exhibit constitutive MLS_B_ (macrolide-lincosamine-type B streptogramin) resistance[25]. Point mutations in 23S rDNA at *E. coli*-equivalent bases 2058(A>T, A>G) and 2059(A>G), which affects the peptidyl transferase region of the ribosome, makes *P. acnes* exhibit constitutive MLS_B_ resistance, and macrolide-only resistance, respectively[26].

As the dominant organism of skin follicle microbiota, *P. acnes* takes up approximately 50% to 90% of organisms in skin follicle[27 28], which is a huge advantage of utilizing comedone as the test specimen to interrogate *P. acnes* susceptibility. To eliminate the interference caused by the nucleic acid of other organisms, primers were designed against the sites that showed greatest inter-species variation. Sequences of ACQUIRE primers and probes were shown in Figure 2. The concentration of is.

**Figure 2.**
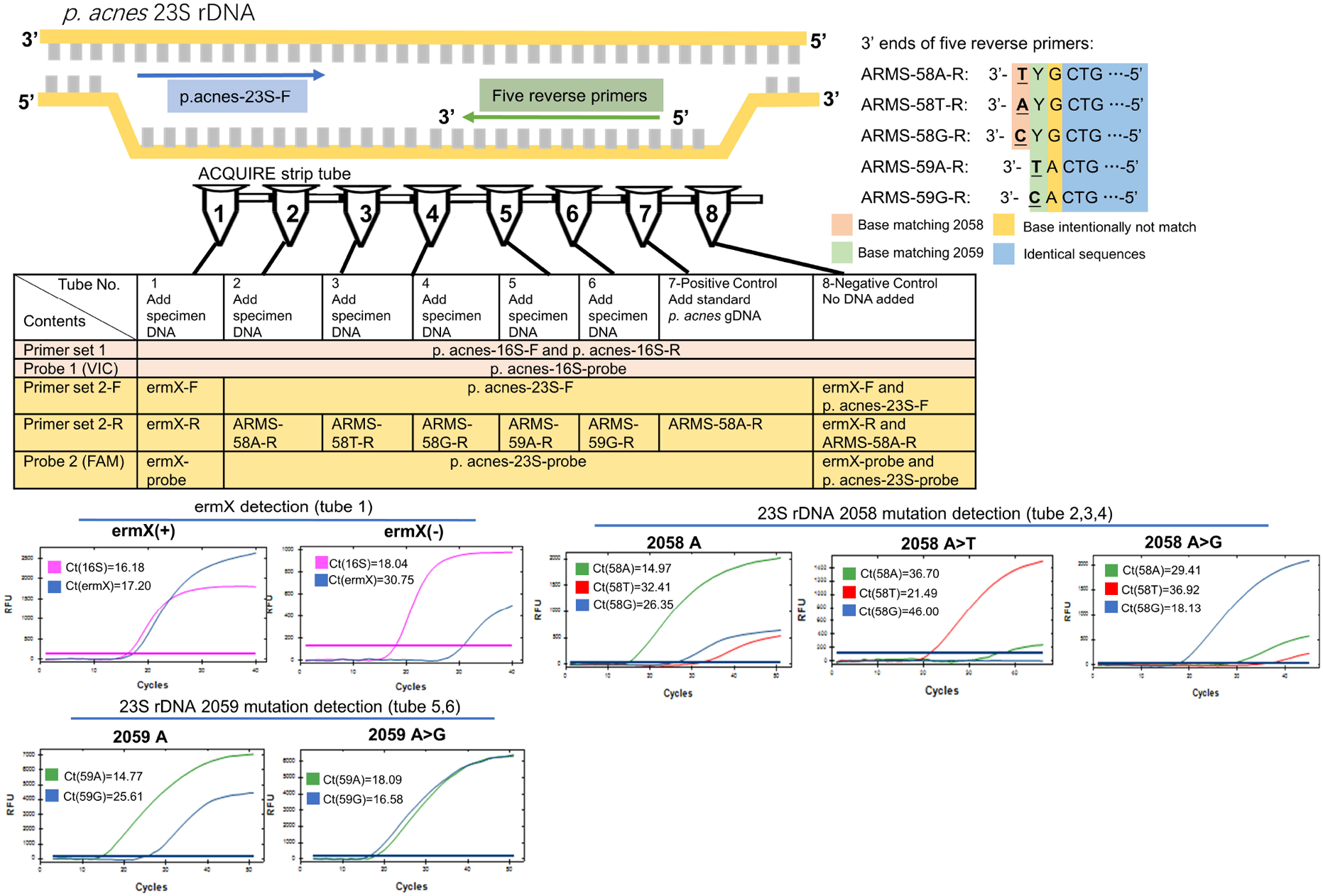
Working principle, composition and typical results of ACQUIRE. Composed of 10 PCR primers and 3 fluorescence probes, ACQUIRE is integrated into one 8-strip tube. Tube 1 interrogates ermX presence, tube 2,3, discriminates 2058 A>T and 2058 A>G from 2058A. Tube5 and 6 discriminates 2059 A>G from 2059A. Tube 7 and 8 serve as positive and negative control, respectively. The discrimination of 23S rDNA mutations was achieved by the Ct value difference caused by the match or mismatch of 3’ ends of five reverse primers. Additional mutations (base intentionally not match) were introduced into primers to strengthen this effect.

Once the specimen was obtained, it was transferred into a 1.5mL microcentrifuge tube prefilled with 175μL lysis buffer (Cat. 939016, Qiagen) and glass beads (40 mesh, 0.425mm in diameter). Then, the microcentrifuge tube was fixed on a tissue disruptor (4000rpm, 10 mins) to release the *P. acnes* resided inside the specimen. Then, the total bacterial DNA was extracted using a universal gram-positive bacteria DNA extraction kit (Cat. FP211, Tiangen Biotech, China), and dissolved in 100μL sterile TE buffer (Cat. 17000-10, Qiagen). The eluted DNA was loaded into an ACQUIRE strip tube(0.5μL eluted DNA per tube) prefilled with primers, probes, water, and qPCR reaction mix(Figure 2). Then, the qPCR protocol was run and the raw results were analyzed and the result was determined (Figure 2).

### Statistical analysis

Statistical analysis was performed on SPSS 22.0 software (IBM, Armonk, NY, USA). Continuous variables were expressed as mean ± standard deviation (SD) and compared using the Mann-Whitney U test for two groups. Categorical variables were expressed as frequencies and percentages and compared using the chi-squared test (n>5) or Fisher’s exact test (n<5). For all statistical analyses, a 2-sided *P* < 0.05 was accepted as statistically significant.

## Results

### 1 ACQUIRE results were as accurate as sequencing method, but cost much less time

Among 106 subjects, the results of ACQUIRE and PCR +Sequencing method were not significantly different concerning the macrolide susceptibility phenotype (*P* = 1.000) (table 1). In ACQUIRE results, 31 (29.2%) subjects were macrolide-susceptible genotype and 75 (70.8%) were macrolide-resistant genotype. In the results of PCR +Sequencing method, consistent with ACQUIRE results, 31 (29.2%) subjects were macrolide-susceptible genotype and 75 (70.8%) were macrolide-resistant genotype (table1). Compared with ACQUIRE, PCR +Sequencing method, which is culture-dependent, cost much more time (Figure 3).

**Table 1.**
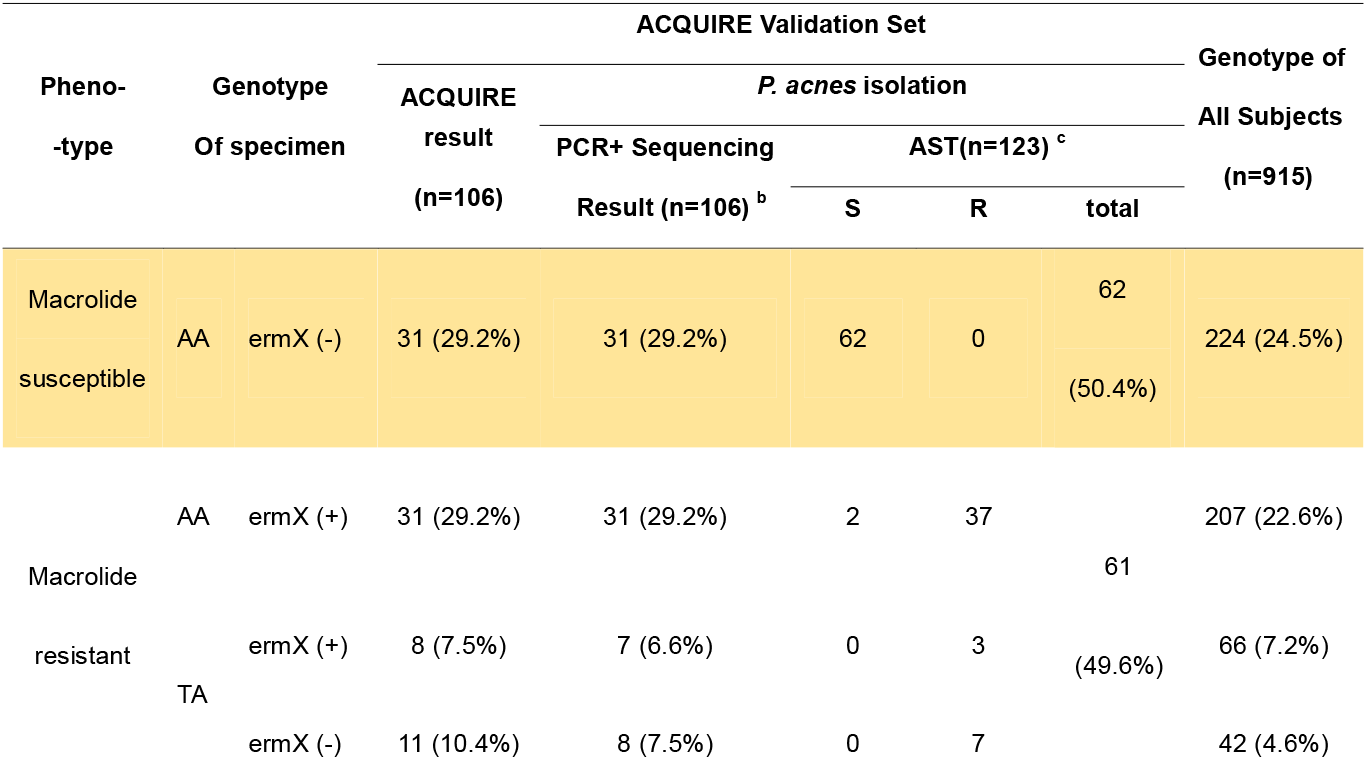
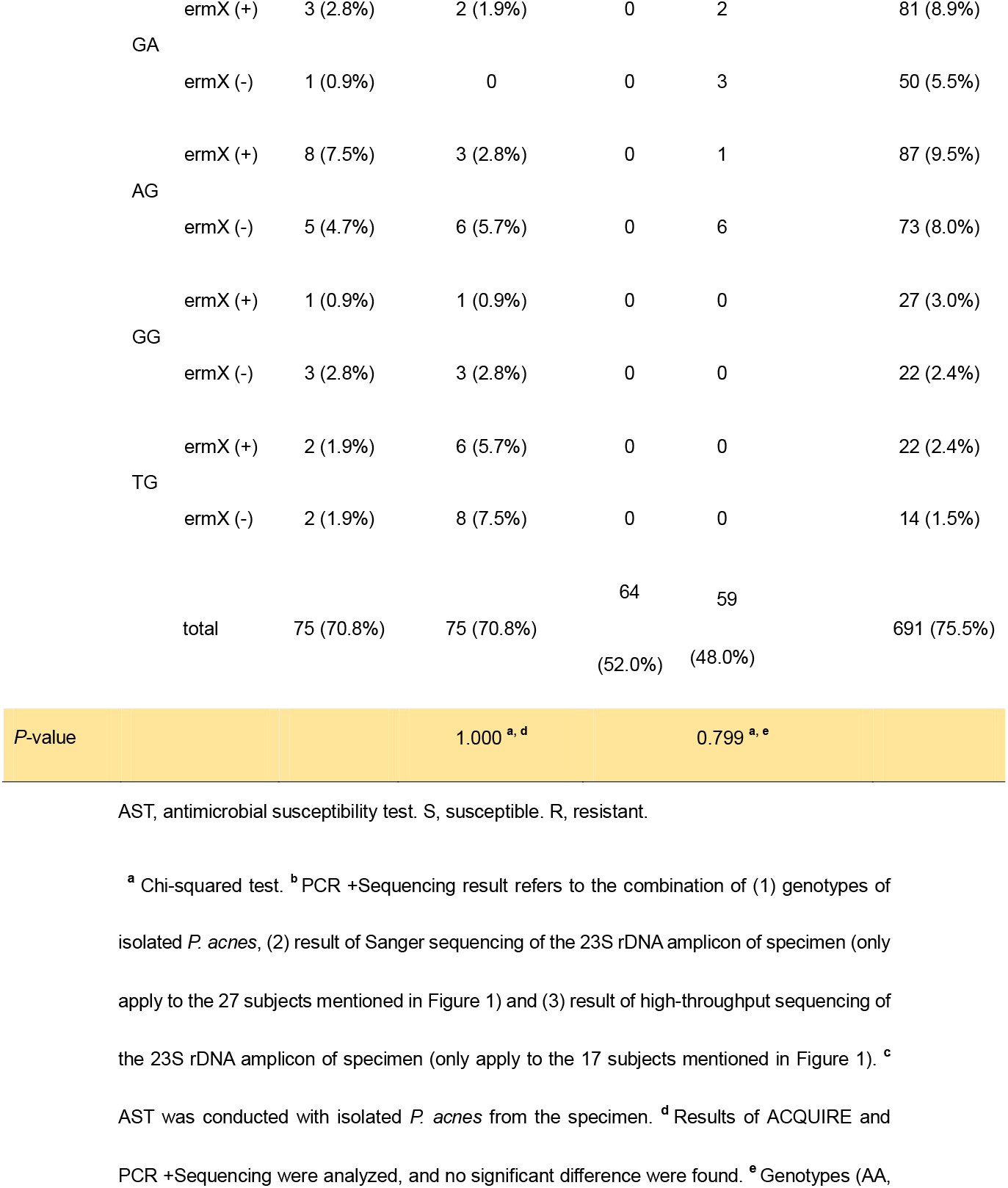
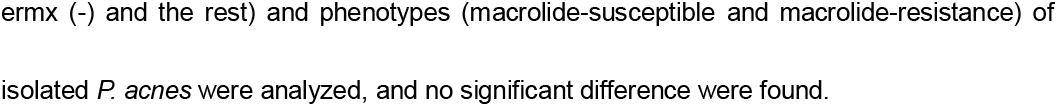
ACQUIRE is a sensitive and accurate method for detecting *P. acnes* resistance.

**Figure 3.**
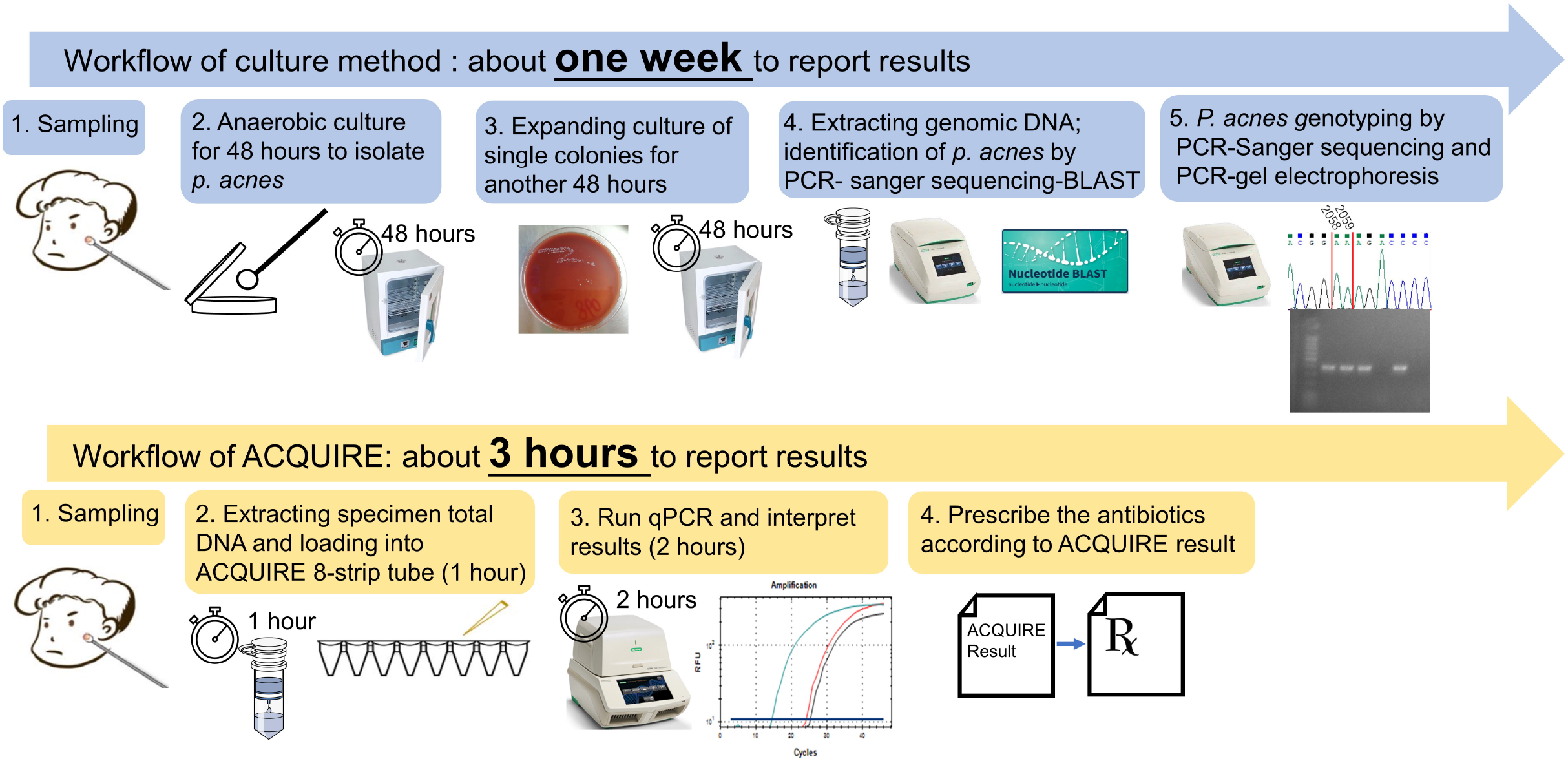
Comparison of the workflow and time-cost between the current method and proposed ACQUIRE.

### 2 Erythromycin susceptibility was consistent with genotypes of *P. acnes*

Total of 123 *P. acnes* strains were isolated from 106 subjects, and their genotype and erythromycin susceptibility were determined. Consistent with previous studies, the genotype of *P. acnes* well correlated with their phenotype of macrolide susceptibility (*P*=0.799), with only two inconsistent strains (table 1).

### 3 ACQUIRE revealed a higher macrolide resistance rate due to more prevalent 23S rDNA mutation

Among 915 subjects whose specimen was tested by ACQUIRE, ermX was the most frequently detected macrolide-resistance determinant and was discovered on 472 (53%) subjects, which concurs with previous studies[14]. However, the prevalence of 23S rDNA mutation is much higher than previous reports, with 468 (52%) subjects carried at least one mutation. The proportion of macrolide-resistant subjects was also higher, with 691 (75.5%) was resistant to macrolide (table 1).

## Discussion

The increasing *P. acnes* resistance asks for dermatologists to utilize those antibiotics that are still effective, which is a limited and diminishing clinical asset, more prudently and smartly. However, due to the inability of the current culture-based method, the status quo of *P. acnes* resistance is underestimated, and the related clinical and microbiological studies are insufficient.

Simply abolishing macrolide and initiating the extensive use of tetracyclines will result in the rapid escalation of tetracycline resistance and cause a waste of antibiotic resources that are insufficient already. Therefore, the most desirable practice for dermatologists would be prescribing tetracyclines to those patients who carry macrolide-resistant P. acnes, and administering macrolide to those whose P. acnes is susceptible to macrolide.

Over the last twenty years, multiple studies[12 16 21 24-26] have well established and confirmed the correlation between the *P. acnes* genotype (23S rDNA 2058_2059 mutation and ermX carriage status) and its macrolide susceptibility phenotype, which laid the foundation for developing a macrolide resistance assay for *P. acnes* based on its genotype.

As a supplement to the current acne treatment algorithm, ACQUIRE can be used whenever a macrolide agent is intended to be prescribed, including topical and oral agents, and leave the rest of treatment options uninfluenced, which made it easy to integrate ACQUIRE into current treatment modalities and guidelines. ACQUIRE also provides an easy, scalable and economical way to probe into the *P. acnes* resistance status in a large population.

By eliminating antibiotic misuse, ACQUIRE can make acne treatment more effective with less cost and shortened course of the disease, representing an opportunity to improve the quality of care. In this real-world study, we demonstrated the possibility and efficacy of integrating ACQUIRE into acne treatment modalities and achieved the better clinical outcome.

In the future, with advancements in the field, functions of ACQUIRE can be further extended, for example, by adding more resistance determinants into the current assay panel to enable the antibiotic resistance detection of tetracyclines and other antibiotics, or enabling the discrimination and quantification of different *P. acnes* subgroups.

Meanwhile, general principles to slow the development of antibiotic resistance should always be encouraged, like using BPO, strictly limit long-term (more than 2 months) antibiotic usage[29] and adopting alternative therapies like laser and light-based treatments.[30]

However, this study has some shortcomings. As a real-world study, the sample size in the Precision Treatment Set is relatively small, and the study was limited by the short duration of the treatment and follow-up periods, which did not address the long-term treatment efficacy. Additional large-scale multi-center randomized controlled trials with longer follow-up time are urgently needed in order to provide more solid evidence for the application of ACQUIRE.

## Conclusions

Macrolide resistance of *P. acnes* is higher than previously reported due to more prevalent 23S rDNA mutations. Future studies should focus on the long-term efficacy of treatment guided by ACQUIRE in large-scale cohorts.

## Data Availability

available upon request

## Notes

Funding sources: China Hunan Provincial Science and Technology Plan (2018SK7005, 2017SK2092); Innovative Education Reform Program of Central South University (2019CG045); National Undergraduate Innovation Training Program of Central South University (201810533233, GS201910533150X); Hunan Science and Technology Innovation Talent Program (To Yiming Xu).

Conflicts of Interest: Feizhou Zhu, Jianglin Zhang, Liyu Chen, Jingheng Zhang and Fang Yu reported China Patent 2020101633011, a method that promotes the detection accuracy and efficiency of P. acnes antibiotic resistance.

### Competing Interest Statement

Feizhou Zhu, Jianglin Zhang, Liyu Chen, Jingheng Zhang and Fang Yu reported China Patent 2020101633011, a method that promotes the detection accuracy and efficiency of P. acnes antibiotic resistance.

### Funding Statement

China Hunan Provincial Science and Technology Plan (2018SK7005, 2017SK2092); Innovative Education Reform Program of Central South University (2019CG045); National Undergraduate Innovation Training Program of Central South University (201810533233, GS201910533150X); Hunan Science and Technology Innovation Talent Program (To Yiming Xu).

### Author Declarations

IRB approval status: This study is approved by the Ethical Committee of Central South University Xiangya Hospital (No.201801002).

